# Use of community-based intervention to promote Family Planning use among Pastoralist women in Ethiopia: Cluster randomized controlled trial

**DOI:** 10.1101/2020.05.29.20117093

**Authors:** Mussie Alemayehu, Araya Abrha Medhanyie, Elizabeth Reed, Afework Mulugeta Bezabih

## Abstract

**Objective:** We assessed the effect of a community-based intervention for promoting FP use and intention among pastoralist communities of Afar region state, Ethiopia.

**Methods:** The design was parallel, cluster randomized controlled trial (CRT) recruiting married women. It had three arms: 1) women’s FP education, 2) male involvement and 3) control with one to one ratio. A total of 33 kebeles were randomized and allocated. Data were collected through an open data kit. Women’s FP education and male involvement in FP service were the interventions. It was implemented with the assistance of a faema leader using a separate group meeting for males and women and disseminates video recorded messages on FP. The intervention was given for a total of 9 months. FP use and intentions were measured. Cluster level summaries considering a cluster effect analysis was performed. The result was presented with adjusted risks and 95% CI. A p-value < 0.05 was used to declare statistically significant.

**Results:** There was a positive change in the proportion of married women who use FP in the women’s FP education arm, absolute risk (AR) of 0.13(95% CI,0.08,0.17) and male involvement arm with AR of 0.29 (95% CI, 0.23,0.34) as compared to the control arm. In the control arm, the proportion of FP use was 4.3%, whereas it was 17.5% with women who receive FP education and 34% in the male involvement arm. Furthermore, the proportion of married women who had high intention to use FP was high in arms of women’s FP education and male involvement with AR = 3.4(95% CI: 2.48,4.91) and AR = 2.1 (95% CI: 1.5,2.95), respectively as compared with the control arm.

**Conclusion:** The community-based intervention (male involvement and women’s education about FP use) brings a significant change in increasing FP users and promotes the intention to use FP.

## Introduction

In the developing world, including Ethiopia, maternal and child morbidity and mortality have remained high, despite the need for urgent action as declared by different international agreement {Hosmer, #25}. Such a burden can be averted by using effective family planning (FP) as a means to decrease unintended pregnancy and its consequences and thereby reduce maternal and newborn morbidity and mortality{Hosmer, #25}. Even though in 2016, there were 300 million women and girls from developing countries using FP -{Hosmer, #23}-, a considerable number (214 million) of women still had an unmet need for FP in the same year{Hosmer, #25}. Sub-Saharan African (SSA) including Ethiopia accounts for the highest number of unmet needs for FP among women of reproductive age group with a value of 24.2 and 26%, respectively {Hosmer, #36}. Apart from this, there is a target to have access and use of FP for an additional 120 million women by the year 2020{Hosmer, #23}. In Ethiopia from 2014–2018, the Reproductive, Maternal, and Newborn Health Innovation Fund (RIF) launched a project aimed to enhance the maternal and child health indicators including FP of the pastoralist communities. All these indicators are far away and lower as compared with the national average. As part of the project, we intend to provide evidence with innovative solutions for service with low coverage like FP in the Afar region, Ethiopia (4).

Ethiopia sets a goal to achieve a contraceptive prevalence rate (CPR) of 55% and total fertility rate (TFR) of 3 by the year 2020 -{Hosmer, #1}- from the current CPR of 41%-(6)- and TFR of 4.6{Hosmer, #5}. However, FP use among pastoralist communities’ is lower than the national average. Afar region is one of the pastoralist regions characterized by pastoralist/agro-pastoralist dominating population. The region’s FP utilization was low characterized by CPR ranges from 5.4 –12.7% -{Hosmer, #5;Hosmer, #26;, 2019 #42;Hosmer, #26}-, TFR of 5.5, and unmet need for FP of 17.2% (12.9 for spacing and 4.3% for limiting) {Hosmer, #5}. It should be noted that religious rooted convictions and norms against the use of FP and strong husbands’ objection towards FP are likely to be the main causes of low FP use and large family size. Hence, such kind of negative norms on FP overshadows women’s decision making power in their families and limits their right to access the FP service {Hosmer, #2;Hosmer, #4}.

Current evidence suggests that educating women on FP and male involvement in FP education based on our previous assessment on best practices for addressing socio-cultural barriers related to FP (11). In addition to this, studies done elsewhere outside the pastoralist community shows an increment of FP use using different modalities. For instance, a study in different districts of India deploys an intervention like having group meetings, providing training to rural providers and community leaders -{Hosmer, #15}- creating awareness and encouraging inter-spousal communication to enhance FP use{Hosmer, #16}. Importantly, previous studies in the pastoralist community had already brought a satisfactory change in promoting maternal, newborn and child health (MNCH). For instance, male involvement, one health approach, having a migratory route of container clinic, mobile clinic and building maternity waiting for home are some of the interventions that made a significant contribution in escalating MNCH {Hosmer, #40;Hosmer, #41;Hosmer, #29;Hosmer, #30}. In addition, a study done in a pastoralist community of Kenya to promote male involvement in the MNCH service utilization and to share with other community members under their catchment had brought an improvement in the MNCH service utilization {Hosmer, #29}. Along with, evidence shows that as the women’s decision-making capacity increases, the proportion of women who use FP increases {Hosmer, #5;Hosmer, #28}. Women’s decision making regarding FP can be further enhanced through involving a male partner in FP service {Hosmer, #33}. Therefore, women’s education and male involvement can be done at the community level to increase the low FP use in the Pastoralist community. In Afar, there is a traditional community structure that serves as a social support group and has high community acceptance called “Faema”{Hosmer, #9}. Moreover, we have a dearth of evidence on quantifying the effect of a community-based intervention (women’s education on FP and male involvement) for enhancing the number of FP use and intention among married women in the pastoralist community.

Furthermore, the Health Extension Program (HEP) which is believed to be one of Ethiopia’s best programs in disease prevention, promoting good health and the use of FP service is less practiced in Afar {Hosmer, #6;Hosmer, #7}. In addition, the women development army (WDA) that was established for the purpose of strengthening the HEP in creating awareness, increase health-seeking behavior and building a community sense of ownership has not been yet established despite its practice in the agrarian region of the country{Hosmer, #7;Hosmer, #8}. Hence, using the community-based intervention with a focus of approaching the community with the community: male to male and female to female was used. Faema has had a good community acceptance, has a separate structure for male and female, with a very long history, and is suitable and feasible for enhancing FP use and intention. Therefore, we hypothesized that educating women on FP and having male involvement on FP education to increase the number of married women who use FP and intention using rigorous method (cluster randomized controlled trial(CRT)) which is practically feasible, cheap and prevents contamination of the disseminated information at the ground (24). The main aim of the current study was to implement and evaluate the effect of a community-based intervention (women’s education on FP and male involvement) using CRT on FP use and intention among married women in Pastoralist community of Afar region, Ethiopia

## Methods

### Design

A cluster-randomized trial of parallel design with three arms (women’s education in FP, male partner FP education and control) was used. One to one ratio allocation of the intervention with a control arm was employed to assess the effect of a community-based intervention in increasing family planning (FP) use and intention among the pastoralist community. The community-based interventions were women’s education in FP and male involvement using faema leader to FP use and intention. Hence, each intervention was given separately for males and females and comparison was made the control arm against women’s education in FP and male involvement arms. The intervention was given at the center of the cluster with 9 months duration. A total of 18 sessions of education for women and men was delivered every two weeks for one hour so as to enhance the FP use and intention.

### Participants

The clusters included in the study had at least 30 households with married women. Inclusion criteria for individuals were: being married, resides in a given cluster as usual place and volunteer to participate, whereas those who declared infertile and seriously ill during data collection were excluded.

### Study setting and period

The cluster randomized controlled trial was conducted in the Afar region. Afar Region is one of nine regional states of Ethiopia. The region is composed of five 5 zones, 32 districts, five town administrations and 404 kebeles (sub-districts), and having an estimated population of 1,816,304 out of those 799,174(44%) are females. Majority of the population reside in rural, and are pastoralists or agro-pastoralist in occupation and are Muslim religion followers {Hosmer, #1}. Three districts namely Mille, Afambo and Kori were included in the intervention. The region is characterized by high early marriage which is mainly influenced by parental decision, and with a high prevalence of early pregnancy and delivery. It also expressed with high illiterate rate, high TFR, low CPR and high unmet need for FP {Hosmer, #5;Hosmer, #3;Hosmer, #6}. A clan-based system favoring large family size, being in a polygamous union of marriage, and a high burden of work among the women is a peculiar characteristic of the Afar women {Hosmer, #10;Hosmer, #8;Hosmer, #9}. Poor access to health care forces women to travel long distances and often demand the accompany of family members to seek health care including FP. The intervention was carried out for 9 months; from January to September, 2018.

### Interventions

The intervention targeted at the cluster level. An integrated behavioral model (IBM) was used to guide the intervention {Hosmer, #27}. The community-based intervention was done to increase the number of women who use FP and intention. These interventions were educating women about FP and involving a male partner in FP education. The approach was designed with the principle of approaching the community with their community member/faema leaders. Initially, intensive training was given for the faema leader on a different aspect of FP by the research team. We trained 2 female and 2 male faema leaders from each cluster for 03 days. In addition to the provision of health education at the cluster level, the health care providers working in FP at the intervention arms took orientation and training on making the health facility ready for the service, availing method mix, managing side effect, and counseling married women on FP use based on informed consent. Furthermore, the HEW at the intervention cluster were trained on how to assist faema leader during FP related health education programs and provide house to house counseling to voluntary married women on how to use FP services. They also facilitate opportunities for using health centers when married women prefer to use a long-acting contraceptive. It should be noted that, before we provide the FP message to the community a tailored message which is highly acceptable in the community was discussed. Accordingly, the emphasis of the message was given to spacing than limiting the number of children.

#### Women’s Education on FP use

The women’s education on FP use was given by female faema leader. Faema is a traditional community structure that serves as a social support group {Hosmer, #9}. Initially, intensive training was given for the faema leader about FP by the research team. The training includes a detail description of Muslim dominating countries’ FP experience and its relation with TFR and maternal mortality –(28)- and starts positively influencing the neighbors and women’s in their catchment on how to use of FP. After the training, with the mobilization of the faema leader, a regular meeting on FP was organized at the center of the cluster. The meeting was held twice a month with a 1-hour duration and it was done in the afternoon. The intervention was given for a total of 9-months. A constant schedule was prepared so as to keep the provision of health education message uniform across clusters in each session. The content of the sessions included information related to the definition of FP, type of FP, the purpose of FP, effectiveness, and duration of prevention. It also included sessions that covered myths and misconception on FP and its side effects, how to overcome the pressure/ resistance comes from influential groups (husband, neighbors, clan and religious leaders) on FP and being a role model by starting using of FP (Figure 1). A logbook or registration was prepared to follow the progress of the intervention. It contains the name of the participants, age, and type of topic discussed FP in each session of health education. The logbook was checked for its delivery by the research team once a month. In addition to educating the women on FP, we use a video recorded message that deals on the district’s FP experts’ experience and information from those married who had already started using FP service were disseminated to the community. The video message from married women deals with the life experience related to FP (its process, benefit, possible challenges, and action was taken). And the video from the FP expert includes the benefit of FP, type of FP, possible side effect and its management of FP. The recorded video message on FP was uploaded to tablet smartphone. The tablet with its accessories was given for the faema leader to disseminated the FP message to the women under their cluster. Training on how to operate, deliver and teach the recorded video message was demonstrated and remonstrated by the faema leader. All the FP message was prepared in the local language “Afarri”.

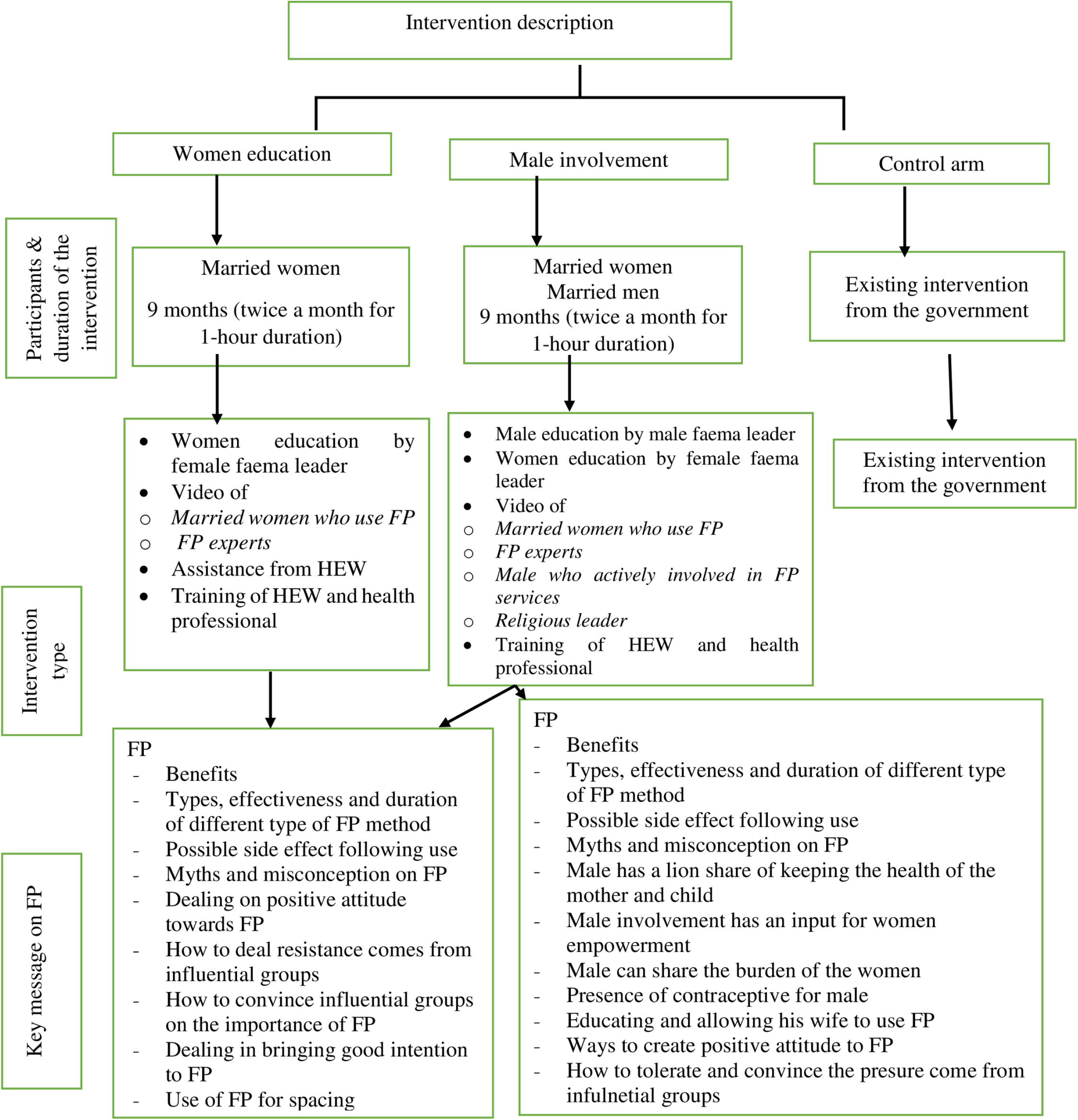

#### Male involvement in FP service

It provides an FP education message for the male partner and married women by the male and female faema leader, respectively. Intensive training was given to the female and male faema leader. The schedule, content, and contact health education hours for married women in the male arm was similar to the women’s education on FP arm. For the male in this arm, the schedule and time allocated for health education were similar to the faema leader, however, the content of the health education gives due focus on the active involvement of males in FP service. It includes allowing his wife to use FP, accompanying her to the health facility, reminding her of the schedule of taking FP, participating in making choice on the type of FP, providing her financial support and helping her in domestic activity. Analogous to the women’s education on FP arm a video recorded message on FP was used to provide health education messages. These video messages were recorded from married women who started using FP, a male who actively involved in FP services, religious leaders and FP experts. The video message from women who started FP use and FP expert was in line with women’s education on FP arm (Figure 1). In the beginning, the importance of FP was discussed with the religious leader and consensus was reached. After the religious leader agreed on the importance of FP, the message on FP vs Islamic religion with the focus of FP use did not contradict their religion. And these messages were recorded and used to teach the male in the male involvement arm. Moreover, the life experience of those men who actively involved in FP services such as; allowing his wife to use FP, accompanying her to health facility, participating in choosing the type of FP, providing financial support and helping her in domestic activity was recorded and used to teach the male in the male involvement arm.

### Measurement of the outcome variables

The purpose of this study was to evaluate the effect of community-based interventions (women’s education on FP and male involvement in FP services) compared to the control group at the cluster level to increase the women who use FP and intend to use. The primary outcome was FP use with the question of “*Are you or your partner currently doing something or using any method to delay or prevent getting pregnant*”. Moreover, the type of FP currently practiced was also collected. Intention to use of FP was measured by 9 items with ascending order from the Gutt-man scale {Hosmer, #11} as a secondary outcome. Some of the items of intention included in this study were “*I am not clear with the benefits of using FP, I am willing to use FP to space the number of children and It is expected that women in our community should use FP and so do I*. A 3-point Likert scale was used with the following responses: agree, neutral and disagree. It was categorized based on the response of married women mean value in to “*low intention to use FP*” and “*high intention to use FP*” for those married women who scored mean and below mean and above means, respectively. In addition to the primary and secondary outcomes, the following variables were collected. Heard of FP was measured when the respondent ever heard of FP. The community responsibility was collected to describe the responsibility of her husband either as a clan, religious and faema leader. In line with this, being a faema leader includes the responsibility of married women in the community. Along with a positive/yes response for the current use of FP, the status of her husband to know for the current use of FP and the type of support obtained from her husband included in our study. To list the type of support; accompany the health facility, reminding the schedule for taking the FP, participating in making choice on the type of FP and either helping them in domestic activity or not.

### Any changes to the trail outcomes after trial commenced

Even though, we strictly follow the protocol we have the following deviation from the original: First, at the beginning, we intend to provide the intervention for six months we try to give the intervention for 9 months with the same frequency (twice a month) since the project time extended; second) we plan to analyze that data using GEE which allows for baseline or covariate adjustment in the final model. However, we are unable to run the model with GEE due to the limited number of clusters per arm (< 15). Hence, we use cluster-level summarizes to report our result; Third) we made some adjustments on the schedule and content of the education message based on the request of the participants.

### Study design, sample size determination, and sampling procedure

The sample size was calculated using the literature of Richard and Lawrence-{Hosmer, #12}- to determine the number of clusters required to detect a difference among different arms. Given a current FP utilization in Afar region of 11.6% -{Hosmer, #5}-expected changes to be acquired following the intervention of 20%, 90% power, 95% confidence interval, considering the intracluster correlation of ρ = 0.05, adjusting for non-response of the individual in a household of 20% and a design effect of 2.2. Taking an assumption of an equal number of clusters and the cluster sample size, the final sample size was 33 clusters and 891 married women. One cluster had 27 married women. Per arm, we include 11 clusters and 297 married women. A systematic sampling technique was used to select 27 married women from one cluster. A sampling fraction was calculated based on the total number of married women in the cluster. A random start number was selected to identify the first married woman in the clockwise direction. Hence, 9 clusters (5 male involvement and 4 women education) of Afambo, 7 clusters (5 male involvement and 2 women education) of Mille and 6 clusters (2 male involvement and 4 women education) of Kori were included in the intervention.

### Randomization

We used a cluster randomized controlled trial parallel-group design with three arms. Using a computer-generated random number, the number of clusters was allocated into three arms (women’s FP education, male involvement, and control) in simple randomization. To avoid bias during the process, the allocation of the clusters was done by another researcher and the result was communicated with the principal investigator. Moreover, the participants were blinded to the intervention.

### Data collection tool and procedure

We develop the quantitative tool based on the previous finding that aims to explore barriers and facilitators to Reproductive Maternal Neonatal Health (RMNH)services including FP{Hosmer, #2;Hosmer, #3;Hosmer, #4;Hosmer, #5}. It contains baseline and end-line data collection with a nine-month duration. The tool was piloted on 10% (118) of the sample and it was pre-tested in 5% (45) married women. Six clinical nurse data collectors and 2 supervisors were used to collecting the data after they got training on the items and how to use mobile-based applications. They were assigned to a different cluster of given districts. The baseline and end-line data were collected using an electronically smartphone-based application open data kit (ODK) for 1 month. Immediately after the data checked for its completeness, it was sent to the Mekelle University (MU) server where the data were accessed and utilized by the research team. A repeated cross-sectional data was used to collect the information from married women.

### Data Quality Control

The data collectors and supervisors were trained. Regular supervision and follow-up were made by supervisors. A reliable and valid tool was used. The paper-based items were transferred to the mobile-based application (ODK) which ensures skip pattern; immediate scanning of the collected tool in the server, friendly to use and avoids cost for paper duplication. The progress of the intervention monitored through the logbook. Intensive training was given for faema leaders, HEW, health care providers, and religious leaders. A tablet-based video FP message was employed. Furthermore, information on the data monitoring and safety, a team from Mekelle University, Samara University, and the Afar regional health Bureau was established. The intends to provide health education message about FP. Accordingly, volunteer married women will go to a health facility and counseled to use contraceptives based on their informed consent at health facilities by the health care providers. The research team takes an effort to minimize the risk and maximize the benefit by following the provision of intervention using the protocol. However, there was no risk reported following the provision of the intervention.

### Statistical Analyses

The data collected using ODK was exported to R software version 3.4.2 for analysis. Intention to treat analysis was used as a framework of analysis. All the analysis was used with a 95% confidence interval (CI) and p-value < 0.05 used to declared statistically significant. Since the number of clusters per arm was 11 per arm, a cluster-level summary was used –- to compare the women’s education and male involvement in the FP use group with the control group. A separate cluster-level summary analysis was done to control arm with the woman’s arm and the male arm with the control arm by considering the cluster effect. Finally, the result of FP use and the intention was described with t-test, df, P-value, mean value of both groups (control and intervention) and adjusted risk with its 95% CI. Moreover, the prevalence ratio (the number FP users at the end line divided to baseline) and odds ratio were calculated for FP use and intention to use

## Results

### Participants and cluster flow

A total of 43 clusters were eligible for the study, out of these 7 clusters did not fulfill the inclusion criteria (30 households, which contains married women and less) and 3 clusters were unable to reach due to the breaking of the bridge due to flood. Hence, 33 clusters were allocated to women’s education, male involvement, and control arm. Hence, the 33 clusters were followed and analyzed. And there was no attrition from clusters. The variance of the cluster for women education arm was 10.03 and 12.29, respectively in the baseline and end-line data (Figure 2). The trial stayed for a total of 9 months; start at Jan-2018 and the trail ends on Sep-2018.

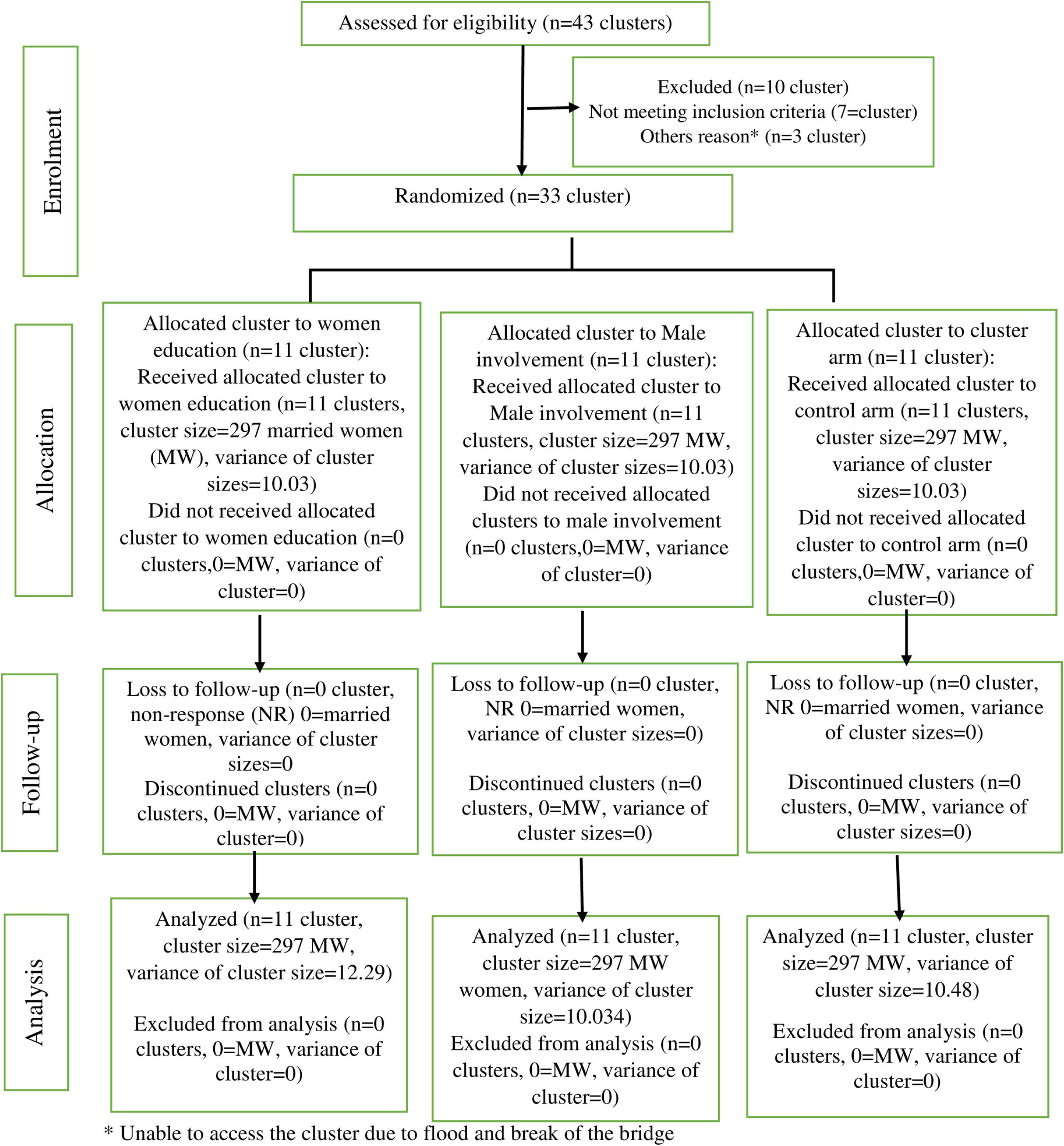

### Baseline and end-line data of respondents

A total of 891 respondents with 297 in each arm participated in the baseline data. In the male involvement arm, the mean age of the respondents was 25.9 (±6. 42), heard of FP 269(90.6) and use of FP 17(5.72). Furthermore, the mean age was 26.8 (±6.10), heard of FP 279(90.3) and use of FP 102(34.3) in the male involvement arm (Table 1).

**Table 1:**
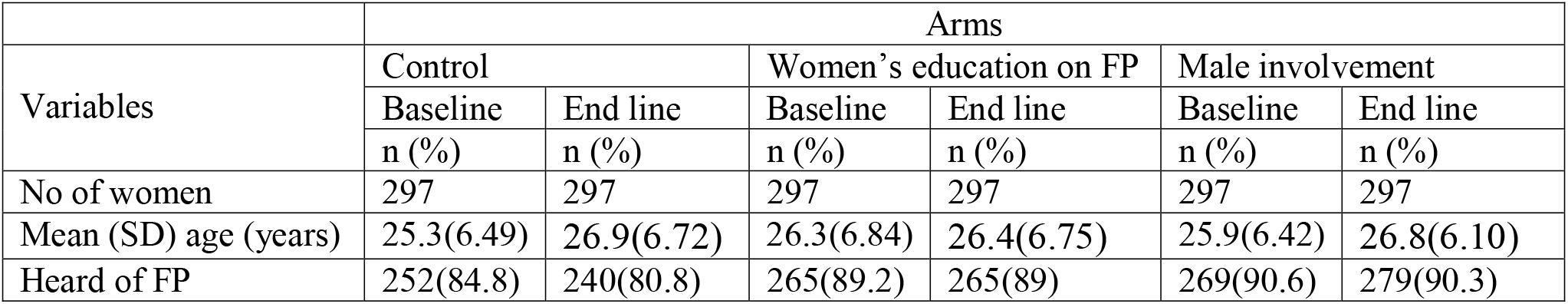
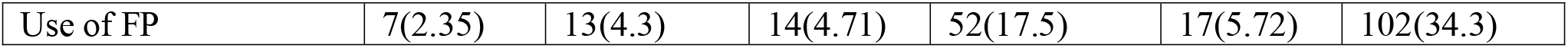
Baseline and end line information in accordance with control, women and male arm among Pastoralist married women Afar, 2019

### Estimation of FP use among married women in accordance with their arms

The level of analysis in this trial was cluster-based. The number of respondents (proportion) reporting yes in use of family planning was 13(0.48), 52 (1.93) and 102(3.78) among the control, women’s FP education and male involvement arm, respectively. Besides, the cluster mean (SD) for responding yes to family planning use was 0.043(0.03) in control, 0.175(0.05) in women’s FP education and 0.343(0.09) in the male involvement arm (Table 2).

**Table 2:** Cluster level numbers and proportions of Pastoralist mothers reporting yes to overall FP use in women’s FP education, male involvement and control arms following the intervention, Afar, Ethiopia, 2019

| Clusters | Arms |  |  |  |  |  |  |  |  |
| --- | --- | --- | --- | --- | --- | --- | --- | --- | --- |
|  |  | Control (m=11 clusters) |  | Women education on FP (m=11 clusters) |  |  | Male involvement (m=11 clusters) |  |  |
|  | Cluster size | # in clusters reporting yes | Cluster proportion reporting yes | Cluster size | # in clusters reporting yes | Cluster proportion reporting yes | Cluster size | # in clusters reporting yes | Cluster proportion reporting yes |
| 1 | 27 | 1 | 0.04 |  |  |  |  |  |  |
| 2 | 27 | 1 | 0.04 |  |  |  |  |  |  |
| 3 | 27 | 2 | 0.07 |  |  |  |  |  |  |
| 4 | 27 | 2 | 0.07 |  |  |  |  |  |  |
| 5 | 27 | 0 | 0.00 |  |  |  |  |  |  |
| 6 | 27 | 2 | 0.07 |  |  |  |  |  |  |
| 7 | 27 | 1 | 0.04 |  |  |  |  |  |  |
| 8 | 27 | 2 | 0.07 |  |  |  |  |  |  |
| 9 | 27 | 2 | 0.07 |  |  |  |  |  |  |
| 10 | 27 | 0 | 0.00 |  |  |  |  |  |  |
| 11 | 27 | 0 | 0.00 |  |  |  |  |  |  |
| 12 |  |  |  | 27 | 6 | 0.22 |  |  |  |
| 13 |  |  |  | 27 | 4 | 0.15 |  |  |  |
| 14 |  |  |  | 27 | 5 | 0.19 |  |  |  |
| 15 |  |  |  | 27 | 7 | 0.26 |  |  |  |
| 16 |  |  |  | 27 | 5 | 0.19 |  |  |  |
| 17 |  |  |  | 27 | 7 | 0.26 |  |  |  |
| 18 |  |  |  | 27 | 2 | 0.07 |  |  |  |
| 19 |  |  |  | 27 | 5 | 0.19 |  |  |  |
| 20 |  |  |  | 27 | 4 | 0.15 |  |  |  |
| 21 |  |  |  | 27 | 4 | 0.15 |  |  |  |
| 22 |  |  |  | 27 | 3 | 0.11 |  |  |  |
| 23 |  |  |  |  |  |  | 27 | 10 | 0.37 |
| 24 |  |  |  |  |  |  | 27 | 11 | 0.41 |
| 25 |  |  |  |  |  |  | 27 | 13 | 0.48 |
| 26 |  |  |  |  |  |  | 27 | 9 | 0.33 |
| 27 |  |  |  |  |  |  | 27 | 14 | 0.52 |
| 28 |  |  |  |  |  |  | 27 | 9 | 0.33 |
| 29 |  |  |  |  |  |  | 27 | 7 | 0.26 |
| 30 |  |  |  |  |  |  | 27 | 8 | 0.30 |
| 31 |  |  |  |  |  |  | 27 | 6 | 0.22 |
| 32 |  |  |  |  |  |  | 27 | 7 | 0.26 |
| 33 |  |  |  |  |  |  | 27 | 8 | 0.30 |
| Totals | 297 | 13 | 0.48 | 297 | 52 | 1.93 | 297 | 102 | 3.78 |
| Mean (SD) |  |  | 0.043(0.03) |  |  | 0.175(0.05) |  |  | 0.343(0.09) |

### Prevalence ratio on FP use based on the baseline and end-line data in accordance with their arms

The number of respondents who use contraceptives was 7 in the control arm, whereas it was 17 in the male involvement arm, during the baseline data. Besides, the number of contraceptive users increase to 102 in the male involvement arm where it was only increased into 13 in the control arm, after the intervention. Hence, the prevalence ratio (end line FP users divided to baseline FP users) was 1.8 in control, 3.7women’s FP education and 6 in the male involvement arm (Table 3).

**Table 3:** Absolute number of FP users and Prevalence ratio in the baseline and end line in accordance with arms Afar, Ethiopia, 2019.

| Variable | Group in accordance with the absolute number of FP users |  |  |
| --- | --- | --- | --- |
|  | Control | Women's FP education | Male involvement |
| Baseline data on FP use | 7 | 14 | 17 |
| End line data on FP use | 13 | 52 | 102 |
| Prevalence ratio (end line/data) on FP use | 1.8 | 3.7 | 6 |

### Characteristics of FP users by study arm

In the male involvement arm, 9(8.8%),10(9.8%) and 17(5.7%) of the FP users accounted for the respondents or their husbands with community responsibility of religious, clan and faema leaders, respectively. Heard of FP and ever use of FP by the respondent was 99(97.1%) and 98(96.1%), respectively in the male involvement arm. The injection was the most common 87(85.3) type of FP used in the male involvement arm. Eighty-eight (86.3%) and 86(97.7%) of the respondent in the male involvement arm, their husband knows for the current use of FP and they provide different types of support, respectively. These supports were accompanying to health facility 66(76.7%), reminding the schedule 81(94.2%) and participating in making choice on the type of FP (Table 4).

**Table 4:** –Description of FP users by selected variables in accordance with their arms in Afar, Ethiopia,2019.

| Variables | Category | Arms |  |  |
| --- | --- | --- | --- | --- |
|  |  | Control | Women's FP education | Male involvement |
|  |  | Use of FP | Use of FP | Use of FP |
|  |  | n (%) | n (%) | n (%) |
| Community responsibility of respondent's husband | Religious leader | 0(0.0) | 3(5.8) | 9(8.8) |
|  | Clan leader | 3(23.1) | 5(9.6) | 10(9.8) |
|  | Faema leader* | 1(7.7) | 5(9.6) | 17(5.7) |
| Heard of FP | Yes | 11(84.6) | 49(94.2) | 99(97.1) |
| Ever use of FP | Yes | 11(84.6) | 48(92.3) | 98(96.1) |
| Type of current FP use | Pill | 1(7.7) | 4(7.7) | 6(5.9) |
|  | Injection | 8(61.5) | 45(86.5) | 87(85.3) |
|  | Implanon | 2(15.4) | 2(3.8) | 8(7.8) |
|  | Others ** | 2(14.4) | 1(1.9) | 1(1) |
| Husband know for using of FP | Yes | 5(38.4) | 39(75) | 88(86.3) |
| Get support from husband for use of FP | Yes | 4(80) | 39(100) | 86(97.7) |
| Type of support from husband on FP use | Accompany to a health facility | 2(50) | 26(66.7) | 66(76.7) |
|  | Reminding the schedule | 1(25) | 33(84.6) | 81(94.2) |
|  | Participate in making choice on the type of FP | 1(25) | 21(53.8) | 62(72.1) |
|  | Helping in domestic activity | 2(50) | 26(66.7) | 64(74.4) |

\*Applicable for both married women and men; \*\*TBA neighbored hood; \*\*\*Condom, Jaddle

### Effect of community-based intervention (women’s education on FP and Male involvement) on FP use

The difference of FP use in women’s education on FP and control arm is 0.13 and the 95% confidence interval is 0.08 to 0.17. And without the intervention (control arm) the proportion of FP use is about 4.3% and with the intervention of women’s education on FP, it is 17.5%, an absolute risk increases of about 13%, but this might be as little as 8% or as much as 17%. In addition, to the male involvement versus the control arm in FP use, the difference is 0.29 and the 95% confidence interval is 0.23 to 0.34. And without the intervention (control arm) the proportion of FP use is about 4.3% and with the male involvement is 34.3%, an absolute risk increases of about 29%, but this might be as little as 23% or as much as 36%. Conversely, the baseline characteristics of FP use was not significantly differenced the control group with male involvement (t = 1.82, p-value = 0.0895) and women education arm(t = 1.4,p-value = 0.1823) Furthermore, the difference of high intention to use of FP and its 95% CI among the women’s education on FP and male involvement with control arm is 0.18(0.03,0.31) and 0.3(0.17,0.42), respectively. And without intervention, the proportion of women who has high intention to FP use is 12.9%, an absolute risk increase of about 1.8% among women’s education on FP and 3% in involving a male partner in FP education (Table 5). Married women in the male involvement and women’s education on FP have an approximate odds ratio of 11.4(6.23,20.93) and 4.6(2.46,8.71) more likely to use FP as compared with the control group, respectively. Similarly, married women in the male involvement arm and women’s education on FP arm have 3.4(2.48,4.91 and 2.1(1.50,2.95) high intention to use of FP, respectively as compared with the control arm.

**Table 5:** Estimated independent t-test coefficients to show the effect of male involvement, women’s education on FP versus control group on FP use, Afar 2019. *significant at p-value <0.05

| Outcome | Mean Value |  | t-test | df | P-value | Absolute Risk | 95% CI |  |
| --- | --- | --- | --- | --- | --- | --- | --- | --- |
|  | Intervention | Control |  |  |  |  | Lower | Upper |
| **FP use** |  |  |  |  |  |  |  |  |
| Male involvement | 0.34 | 0.043 | 10.01 | 12.346 | 0.0000002* | 0.29 | 0.23 | 0.34 |
| Women's education on FP | 0.17 | 0.043 | 6.59 | 15.74 | 0.000006* | 0.13 | 0.08 | 0.17 |
| **Intention to use of FP** |  |  |  |  |  |  |  |  |
| Male involvement | 1.59 | 1.29 | 5.14 | 19.7 | 0.00005* | 0.3 | 0.17 | 0.42 |
| Women's education on FP | 1.47 | 1.29 | 2.52 | 19.19 | 0.02* | 0.18 | 0.03 | 0.31 |

### Any potential harms of the trails

The study report that there was no adverse event following the provision of the intervention due to the following reason; 1) the decision for taking contraceptive mainly depends on the informed choice of the married women, 2) the provision of counseling was provided by trained health professional at healthcare facilities and it includes a management of potential side effect and action to be taken,3) there was a team which deals with data monitoring safety which was responsible for provision of the intervention based on the protocol.

### Discussion

As a limitation of our trial, the assessor of the outcome measures was not blinded to the type of intervention. Since the number of clusters allocated here small, we employ a cluster-level summary that did not account for covariate or baseline data adjustment in the final model. However, the proportion of women who use FP in the control arm vs male involvement and control arm vs women education is not significantly different at the baseline. The intervention was targeted at a group level and it did not consider a couple of bases. And, the intervention period (9 months with the provision of education twice a month) may be a short time to bring a huge change in FP use. Moreover, we are not able to adjust the baseline or covariate during analysis in the final model. And our study will be generalizability intends to similar pastoralist community but mightn’t to the agrarian community. Furthermore, even though CRT prefers to prevent contamination of information, we did not employ a buffer zone. However, the intervention cluster was separated from control villages by a distance of 20–40 km.

Our finding revealed that women’s FP education and male involvement as a community-based intervention bring a significant change in increasing FP use and intention among the pastoralist community. Our findings will be generalizable to a similar pastoralist community. The study was done among 33 clusters with three arms: women’s education on FP, male involvement and control using a cluster randomized controlled trial. It was accompanied by applying different interventions; delivery of FP message by faema leader and disseminating FP message using video of (role model persons, FP experts, and religious leader) for a total of 9 months, with the frequency of twice a month. The intervention was directed through an integrated behavioral model (IBM). The clustering effect was considered during data analysis.

A pre-post study done in Mali revealed that an increment of FP use. The study employs three groups: community-based contraceptive distribution (CBD), education and control. In the CBD, in assistance of the local chief, each village asked to select an FP promoter (male and women). They were responsible to provide FP education via group meetings, home visit and to sell contraceptives for the same sex. In addition to this, intensive training was given for community health agents and nurses to provide education only. Overall, they found an increment of FP along with the three groups with a higher increment in the CBD. In line with the Mali study, our study uses the same principle of approaching males via males and women via males in an area where the women had poor decision-making power in FP. It should be noted we use a community-based structure (faema leader) which has high acceptance in the community. However, our study revealed women’s FP education and male involvement as a component of community-based intervention brings a significant change in increasing FP use as compared with the control arm. This might be due to our approach employed to reduce the workload of the faema leader to give emphasis on the provision of FP education than to sell contraceptives and collect money. Along with this, we have a difference in the provision of health education about FP on a regular basis (twice a month) and the video assistance message of the influential group was included. In contrast, contraceptive stockout, unable to give the education of FP by the promoters due to sick and not having a regular basis in the provision of education was seen challenges in the Mali study.

In line with our study, different study elsewhere done on community-based intervention using different approaches supports our finding. For example, a study done in rural India increased inter-spousal communication (43 vs 13%-point change) and increased uptake of contraceptives uses (27 vs5%-point change in the intervention and control, respectively) among young married couples with a group meeting and individual counseling by strengthening the capacity of frontline health functionaries. The delivery of the intervention was targeted at spousal level (inter-spousal communication and decision making), family level (by sensitizing family members to early and repeated pregnancies), community-level (advocating young married women health needs and rights among influential community members) and health system{Hosmer, #16}. Moreover, in the Kinshasa study, they recruited community-based distributors (CBDs) to have a group discussion, individual counseling, distribution of contraceptive (condom, pills, and CycleBeads) and arranging referral {Hosmer, #14}. In addition, a study done in Bahir district of India reports that an increment of FP following provision of training to the rural provider and community leaders, group meeting and disseminating message using street theatre and wall painting for a total provision of the intervention 21–27 months. A male and female agent was used to approach the men and women group, respectively in providing information and arranging referral as part of its intervention {Hosmer, #15}. Furthermore, a retrospective pre-post study in Pakistan on Community-based integrated approach to changing women’s FP behavior for the past 24 months found that an increment of 10.7% contraceptive prevalence rate with Sukh’s initiative: create awareness, encourages intra-spousal communication, distribution of contraceptive and arrange a referral for better service {Hosmer, #17}. The overall women empowerment could be positively influenced by women’s education with their ability to convince the influential group (husband, clan and religious leader), positively influence their neighbored women and her child, increase access to information, easily absorb health education messages, critically think and take corrective action. Education including women’s education is a good intervention to improve FP use by increasing the health-seeking behavior and health status of pastoralist women. Besides, it increases their ability for self-determination and access to financial freedom to get quality of care including FP service. And evidence shows that empowered women are more likely to seek and use health services including FP. One component of women empowerment is participating women in decision making in all aspects of health including FP. Hence, such empowerment can be facilitated through the active involvement of men in FP use {Hosmer, #5}. Hereafter, such collective influence would help the mother to create comprehensive knowledge on FP, having a positive attitude which in turn has an effect in increasing the FP uses and intention.

A cluster-randomized controlled trial (CRT) with the Bandebereho couples’ intervention engaged men and their partners in participatory, small group sessions of critical reflection and dialogue. From its 15 sessions on a different topic, FP included as one session with the aim of the description on the benefits of FP, provide information on different contraceptive methods and the value of couple communication. And they found a significant improvement of FP in the intervention as compared with the control group {Hosmer, #44}. A similar CRT in India with the focus of married men and couples brings a significant change in contraceptive use. It uses assessment, dialogue, education, FP Goal Setting & Action Plan and provision of condoms and pill as well as contraceptives using trained village health care providers {Hosmer, #46}. Moreover, a quasi-experiment study in rural Vietnam reports a significant change in increasing the intrauterine device (IUD) utilization between the control and intervention groups. The intervention intended to provide tailored messages and counseling on IUCD for the male to make/allow his wife to use in the intervention group, while no intervention was done in the control group. The intervention was made two round contact in 6-months and it was guided by social cognitive theory(35). In line with these studies, our study finds that a significant increment in FP uses for married women in the male involvement arm as compared with the control arm. It should be noted that male involvement helps not only in accepting a contraceptive but also in its effective use and continuation. One mechanism to improve male involvement could be achieved in the provision of health education messages to improve the beliefs and attitudes of men (36). Importantly, our study delivers the health education message on FP in a group setup using the male faema leader to approach the male. However, the Rwanda study uses a couple of bases in the provision of an FP message which ensures spouse communication even though it depends on the local context. For instance, at the beginning raising the issue of FP was considered as a taboo with the negative influence of influential groups (husband, religious and clan leaders). Conversely, approaching males via male in a group discussion on FP and reaching in consensus with the influential groups on its importance is vital and strictly followed to enhance FP use. This implies using an alternative mechanism based on the local context in the disseminating FP message exerts a strong impact on the current practice of and intention to use.

Furthermore, male involvement in the Maternal, Newborn and Child Health (MNCH) in the pastoralist community of Kenya brings an increment in the service utilization other than FP. Importantly, using the male structure (Boma model), they mobilize community, harmonization, and integration of different community structures, women empowerment and enhancing community participation in health service delivery {Hosmer, #29}. This illustrates male involvement is feasible and brings a remarkable change in the pastoralist context. It should be noted, married women in our study embedded in strong religious and cultural perspectives which favors a large family size, husband objection for not practicing of FP and poor decision-making {Hosmer, #4;Hosmer, #2;Hosmer, #3}. Moreover, all aspects of the health and wellbeing of the pastoralist women strongly affected by religious perspective and belief -{Hosmer, #30}- and the community believed that all health problems are caused by supernatural forces {Hosmer, #31}. And, such strong resistance on FP can be resolved with a continuous discussion with the influential groups on the importance of FP as it was evident in our study. For instance, a religious leader preaches the community to use FP for spacing. As a result, a considerable number of married women whose husband participating in community responsibility of either religious or clan leader start to use FP in our study where. Hence, having male involvement and use of religious leader to disseminating information on FP is crucial as it was evident in this study. Having male involvement in FP service would create a better understanding and positive attitude towards FP use. Hereafter, this would create a golden opportunity for the women to use FP, get psychological, emotional and financial freedom to FP use. This would help to improve the health status of the mother and her child by having optimal birth spacing, mitigate unwanted pregnancy and its consequence, improves the economy of the household and mental satisfaction.

Our study deploys different interventions to bring a significant change in FP use. One of the strategies is to train faema leader-{Hosmer, #9}- to teach the community member in their catchment. The teaching community is a novel strategy to bring a behavior change, however, such strategy would be effective if the change comes from the change agent as it was evident in our study a considerable faema leader use FP. Furthermore, we bring a significant change in increasing the number of married women who use FP in our study. However, when we see the method mix, most of the married women use short-acting contraceptives. Hence, further effort is needed to shift the short-acting FP users to long-acting FP which is effective, cheap, reduce work burden and needs less visit of the married women to the health facility{Hosmer, #35}.

Our study account that there was a significant change in intention to use FP in male involvement and women’s education on FP following the provision of FP education by faema leaders. This also supported by other studies conducted elsewhere {Hosmer, #18;Hosmer, #19;Hosmer, #20;Hosmer, #21;Hosmer, #22}. The effect could be explained by the fact that having good/high intention is an important factor for women to use FP by considering the effect of having comprehensive knowledge and a positive attitude towards FP use. This implies that tailoring the intervention based on the context of the pastoralist community using faema leaders brings a significant change in enhancing the number of women who intend to use. However, this requires further effort to create a sense of ownership by the community and discussing with the governmental officials to consider the existing structure in the provision of future intervention about health and health promotion activities.

A study in Mali identifies the following challenges; community reluctance to accept FP message and use, insufficient funds to purchase contraceptive, unable to cover all the segment of the community in the provision of health education, religious influence with the concept of contraceptive use is prohibited by Islam fear of the community member to included their name in the promoter notebook. To overcome such challenges, increasing the number of the education sessions, arranging a group talk in a break and reassured the community names and personal information would be kept confidential by the promoters{Hosmer, #18}. In addition, weak Interaction with clinical Services, having weak support and supervision of Community-Based Distributors and Recurring Stock-Outs of contraceptive{Hosmer, #14}. In line with these studies, our study deploys the following mechanism to minimize the challenges in the provision of the interventions. First, a tailored message which has high acceptance by the community was discussed. Accordingly, the emphasis was given to FP for spacing than limiting and IBM was used to direct the intervention. Second, influential groups like religious leaders were approached and reach in consensus to disseminate information on FP using recorded video. Third, we provide an education message to disclose of FP use to ensure spousal communication and male support. And we reassure the study participants, no personal identifiers will be used and all the information would be kept confidential. Fifth, adequate preparation on the mobilization of the community member to attend the group FP education was done continuously by the faema leader in assistance with the HEW to arrange a referral to provide direct support. Along with a video uploaded a message on FP was used to teach the community. Six we engage the important stakeholder from the beginning to avail contraceptive, method mix, counseling based on the informed consent and to provide the FP service without a fee.

Our study is a novel study on pastoralist communities in the area where married women confined with strong cultural and religious perspectives with the aim of dealing with an innovative approach like using the existing community structure like faema to enhance the number of FP users. In addition, it employs a cluster randomized controlled trial study design, which is methodologically strong. And it employs an integrated behavioral model that guides the provision of the intervention in an area where the married women embedded in the influence of the husband, religious leader and social norms don’t promote FP use. Moreover, approaching the community with the community’s existing structure like faema which has a separate structure for both sexes, high acceptability, and long history would be vital for its feasibility and scaleup leader. Finally, our study uses a reliable and valid tool with the family planning aspect in the pastoralist context.

### Conclusion

Our finding revealed that women’s education on FP and male involvement brings a significant change in increasing the number of women who use FP and intention. Using the faema structure in the pastoralist community appears to be effective in increasing the number of women FP users.

## Data Availability

Our data will be available upon a reasonable request and researchers would access an anonymized version of the data set either from BMJ open journal or through a direct request to the corresponding author via email.

## Declarations

## Abbreviations

AOR: Adjusted Odds Ratio
AMREF: Africa Medical Research Fund
CPR: Contraceptive Prevalence Rate
CRT: Cluster-Randomized Controlled Trial
FID: Development for International Development
RIF: Reproductive Innovative Fund
EDHS: Ethiopian Demographic Health Survey
FMOH: Federal Ministry of Health
FP: Family Planning
HSTP-: Health Sector Transformation Plan
POM-: Proportional Odds Model
PPOM: Partial Proportional Odds Model
TFR-: Total Fertility Rate
WHO: World Health Organization

## Ethics approval and consent to participate

The study protocol was approved by the Institutional Review Board (IRB) of Mekelle University College of Health Sciences with a reference number of ERC 1435/2018. Permission was obtained from all relevant authorities in the Afar regional health bureau and participating district health offices. In the beginning, community consent was secured from the influential group (religious and clan leader). Verbal consent was secured before conducting the interviews. A one-page consent letter was attached to the cover page of each questionnaire as an information sheet which includes a detail description about the purpose of the study, benefit, and risk of participating in the study, participation is on voluntary basis, the right to withdraw from the study, identification of informant was possible only through specific identification numbers and the privacy and confidentiality of collected information. The trial was registered in a ClinicalTrials.gov with a reference number of NCT03450564.

## Consent for publication

Not applicable.

## Authors’ contributions

MA contributed to the initiation of the study, design, data collection, data analysis, and write up. AAM and AM contributed to the initiation of the study, design, and write up. ER contributed to the interpretation of the findings and write up of the manuscript. All authors read and approved the final manuscript.

## Acknowledgment

We are grateful to thank the data collectors, supervisor and study participants for the successful accomplishment of the study. In addition, we would like to thank the faema leaders, health care providers, and religious leaders participating in the provision of the intervention. Finally, we would like to thank, Afar regional health bureau, Mekelle University, DFID and FMOH.

## Strengths and limitations of this study

- It is a novel study on pastoralist communities in the area where married women confined with strong cultural and religious perspectives with the aim of dealing with an innovative approach like using the existing community structure like faema to enhance the number of FP users.
- It employs a cluster randomized controlled trial study design, which is methodologically strong.
- It uses an integrated behavioral model that guides the provision of the intervention in an area where the married women embedded in the influence of the husband, religious leader and social norms don’t promote FP use.
- Approaching the community with the community’s existing structure like faema which has a separate structure for both sexes, high acceptability, and long history would be vital for its feasibility and scaleup leader.
- The assessor of the outcome measures was not blinded to the type of intervention.
- Since the number of clusters allocated here small, we employ a cluster-level summary that did not account for covariate or baseline data adjustment in the final model.
- The intervention was targeted at a group level and it did not consider a couple of bases.
- The intervention period (9 months with the provision of education twice a month) may be a short time to bring a huge change in FP use.

